# Novel highly divergent SARS-CoV-2 lineage with the Spike substitutions L249S and E484K

**DOI:** 10.1101/2021.03.12.21253000

**Authors:** Katherine Laiton-Donato, Jose A. Usme-Ciro, Carlos Franco-Muñoz, Diego A. Álvarez-Díaz, Hector Alejandro Ruiz-Moreno, Jhonnatan Reales-González, Diego Andrés Prada, Sheryll Corchuelo, Maria T. Herrera-Sepúlveda, Julian Naizaque, Gerardo Santamaría, Magdalena Wiesner, Diana Marcela Walteros, Martha Lucia Ospina Martínez, Marcela Mercado-Reyes

**Affiliations:** Instituto Nacional de Salud, Bogotá, Colombia; Universidad Cooperativa de Colombia, Santa Marta, Colombia

**Author notes:** These authors contributed equally to the paper (Katherine Laiton-Donato, Jose A. Usme-Ciro).

**Keywords:** SARS-CoV-2, lineage, COVID-19, Spike, variant

## Abstract

COVID-19 pandemics has led to genetic diversification of SARS-CoV-2 and the appearance of variants with potential impact in transmissibility and viral escape from acquired immunity. We report a new lineage containing ten distinctive amino acid changes across the genome. Further studies are required for monitoring its epidemiologic impact.

---

COVID-19 continues challenging the health system abroad. After the emergence of SARS-CoV-2 in China in late 2019 and despite the rapid international response once the WHO declared it as a Public Health Emergency of International Concern (PHEIC), the virus rapidly crossed the borders, started autochthonous transmission in every country and spread locally despite the strict lockdown measures (1). The enormous population size of SARS-CoV-2 at the global level and its RNA nature has led to the rapid accumulation of genetic variability as more than 800 lineages (2,3). Some lineages or genetic variants have special attention due to the rapid increase in frequency in some areas (4), abnormally high mutation accumulation across the genome, most amino acid changes affecting the spike protein, evidence for evolutionary convergence of some critical changes and increasing evidence for virus escape to the antibody-mediated immunity (5). As the genomic information is being deposited in public databases, more lineages of special attention are being reported (https://github.com/cov-lineages/pango-designation/issues) and a very high and increasing number of lineages containing the E484K substitution in the Spike protein have been reported to emerge independently at least 68 times and worldwide (Table 2). Further studies are required to assess the functional role of these mutations and to monitor their epidemiologic impact.

**Table 2.**
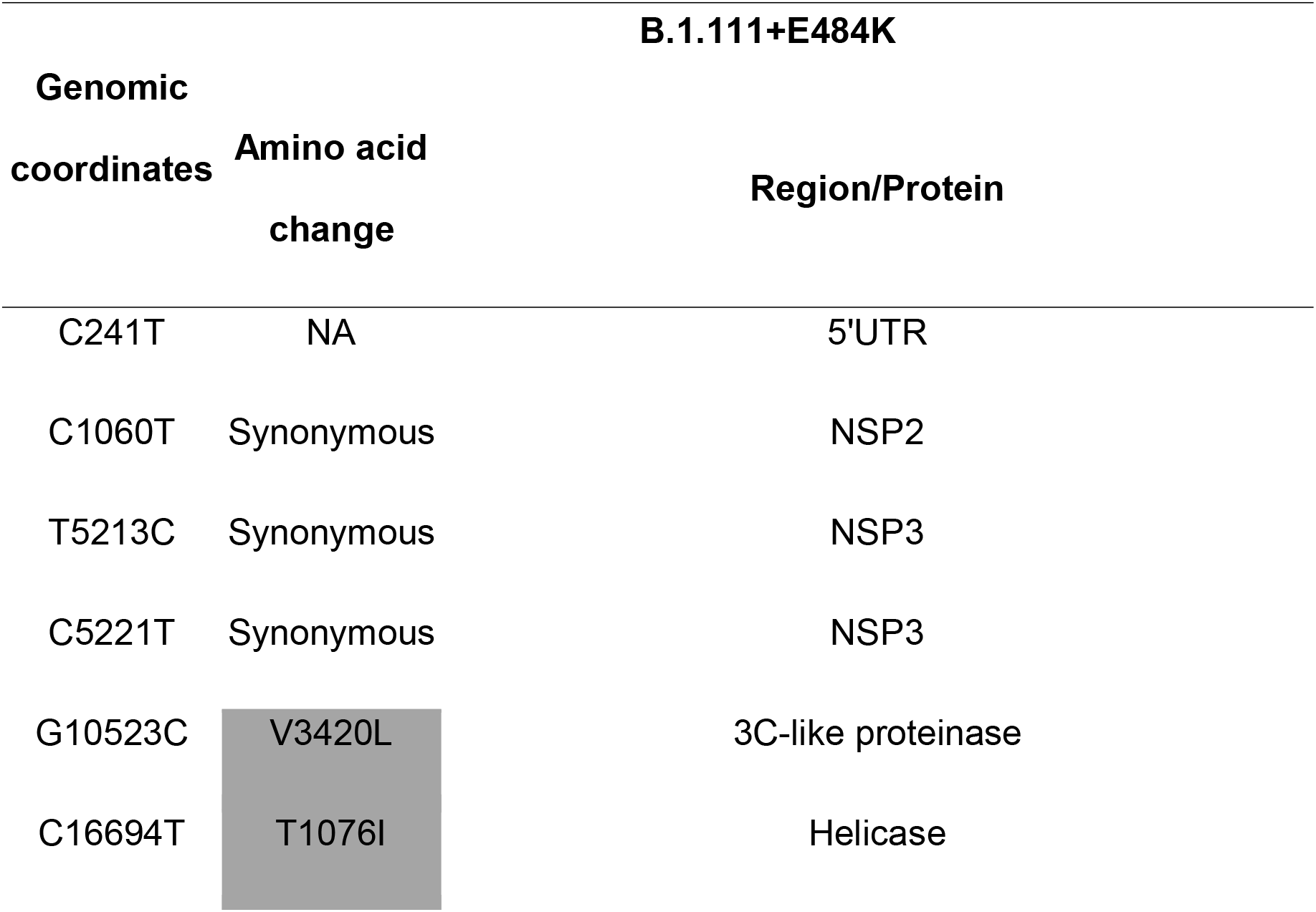
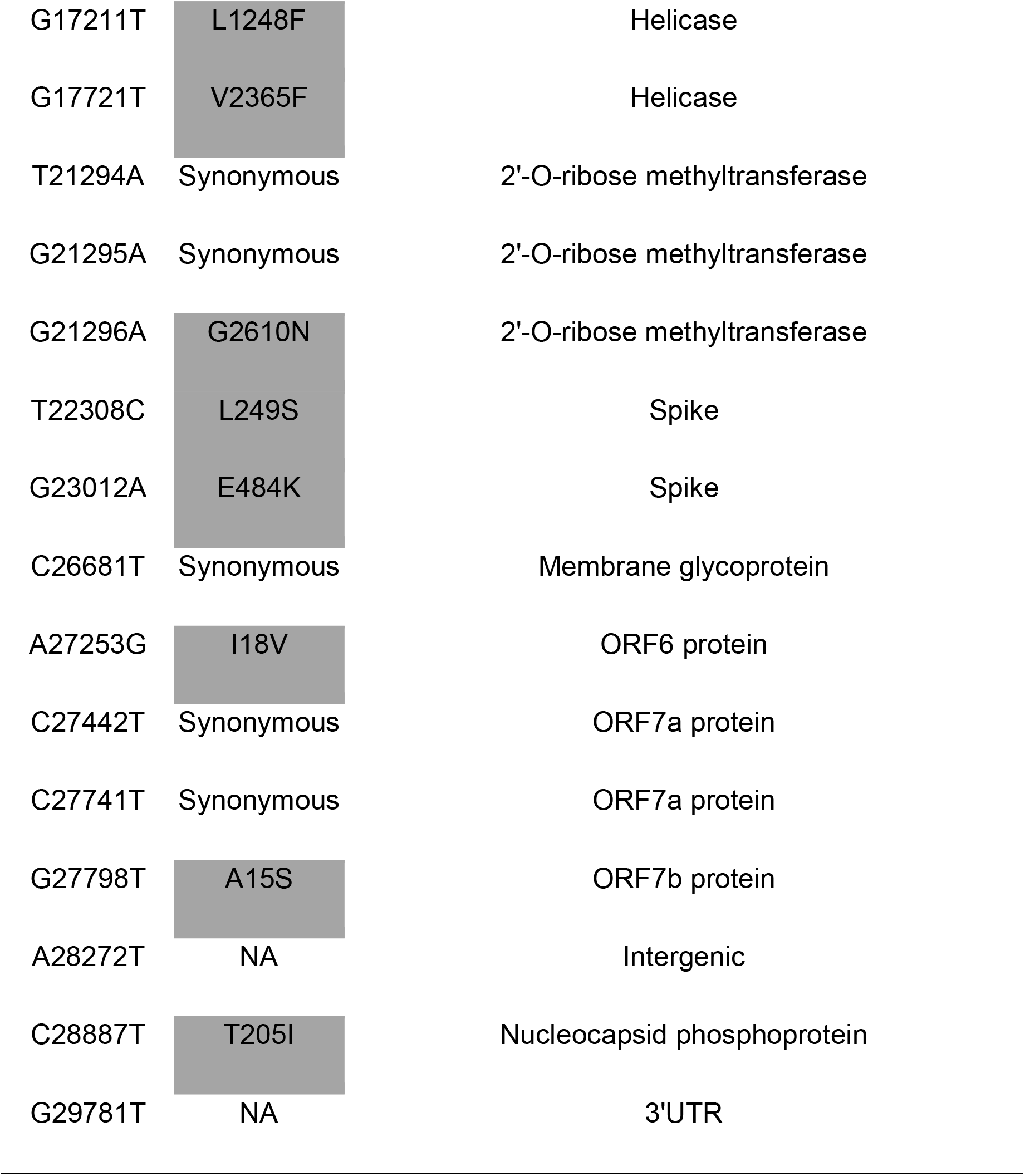
Nucleotide and amino acid substitution pattern of the B.1.111+E484K lineage.

| **B.1.111+E484K** |  |  |
| --- | --- | --- |
| **Genomic coordinates** | **Amino acid change** | **Region/Protein** |
| C241T | NA | 5'UTR |
| C1060T | Synonymous | NSP2 |
| T5213C | Synonymous | NSP3 |
| C5221T | Synonymous | NSP3 |
| G10523C | V3420L | 3C-like proteinase |
| C16694T | T1076I | Helicase |
| G17211T | L1248F | Helicase |
| G17721T | V2365F | Helicase |
| T21294A | Synonymous | 2'-O-ribose methyltransferase |
| G21295A | Synonymous | 2'-O-ribose methyltransferase |
| G21296A | G2610N | 2'-O-ribose methyltransferase |
| T22308C | L249S | Spike |
| G23012A | E484K | Spike |
| C26681T | Synonymous | Membrane glycoprotein |
| A27253G | I18V | ORF6 protein |
| C27442T | Synonymous | ORF7a protein |
| C27741T | Synonymous | ORF7a protein |
| G27798T | A15S | ORF7b protein |
| A28272T | NA | Intergenic |
| C28887T | T205I | Nucleocapsid phosphoprotein |
| G29781T | NA | 3'UTR |

In Colombia, SARS-CoV-2 genomic surveillance was established early during the pandemic, leading to the identification of the importation of at least 12 lineages before international flight cancellation and during lockdown. A 48% of SARS-CoV-2 sequences were assigned the B.1 parental lineage with little or no shared mutations accumulated during the early local transmission inside the country (6). Thereafter, the microevolution of the virus allowed the emergence of some lineages, including the B.1.111 and B.1.420, which are considered Colombian lineages, due to a major representation of sequences from Colombia (37.4% and 85.4%, respectively) in GISAID by February 28, 2021.

Here we report a novel and highly divergent lineage with 21 characteristic mutations, including 10 non-synonymous, 8 synonymous and 3 mutations in non-coding regions (5’and 3’ UTR and intergenic region).

Genomic surveillance was established at the Sequencing and Genomics Group, National Institute of Health, Colombia. Samples for NGS were selected from routine surveillance in all departments and special groups based on clinical and epidemiologic criteria. A total of 287 complete genomes were processed during the period March 2020 to February 2021. Processing of RNA samples was performed as previously described (6), with the suggested modifications of the amplicon sequencing protocol (Arctic LoCost) and NGS raw data processing following the protocol described for ONT. Lineage assignment was performed through the Pangolin algorithm (2). A dataset including Colombian sequences of SARS-CoV-2 representative of the different lineages and those previously reported with substitutions of special interest was used for recombination detection, evolutionary and phylogenetic analyses (Appendix 1).

Four Colombian sequences collected between December 26, 2020 and January 14, 2021with a characteristic mutation pattern, including two amino acid changes in the Spike protein (L249S and E484K) were assigned to the B.1.111 lineage by Pangolin COVID-19 Lineage Assigner (https://pangolin.cog-uk.io/). The lineage B.1.111, first detected in the USA from a sample collected on March 7, 2020 and subsequently in Colombia on March 13, 2020 is currently circulating and mainly represented by Colombian sequences from all around the country (https://microreact.org/project/vHdc5J3MeoYJ2u69PLP6NF). Recombination analysis was performed by RDP4 software with six statistical tests (*P*-value □ < □0.05) and no putative recombination events were found. To reconstruct the phylogenetic relationships of the B.1.111+249S/484K sequences with other SARS-CoV-2 sequences representative from the different lineages circulating in Colombia and to determine their recent origin we estimated a ML phylogeny. These preliminary results are consistent with the emergence of a novel and phylogenetically distant lineage of SARS-CoV-2. Phylogenetic reconstruction (Figure 1) shows the B.1.111 as the parental lineage.

**Figure 1.**
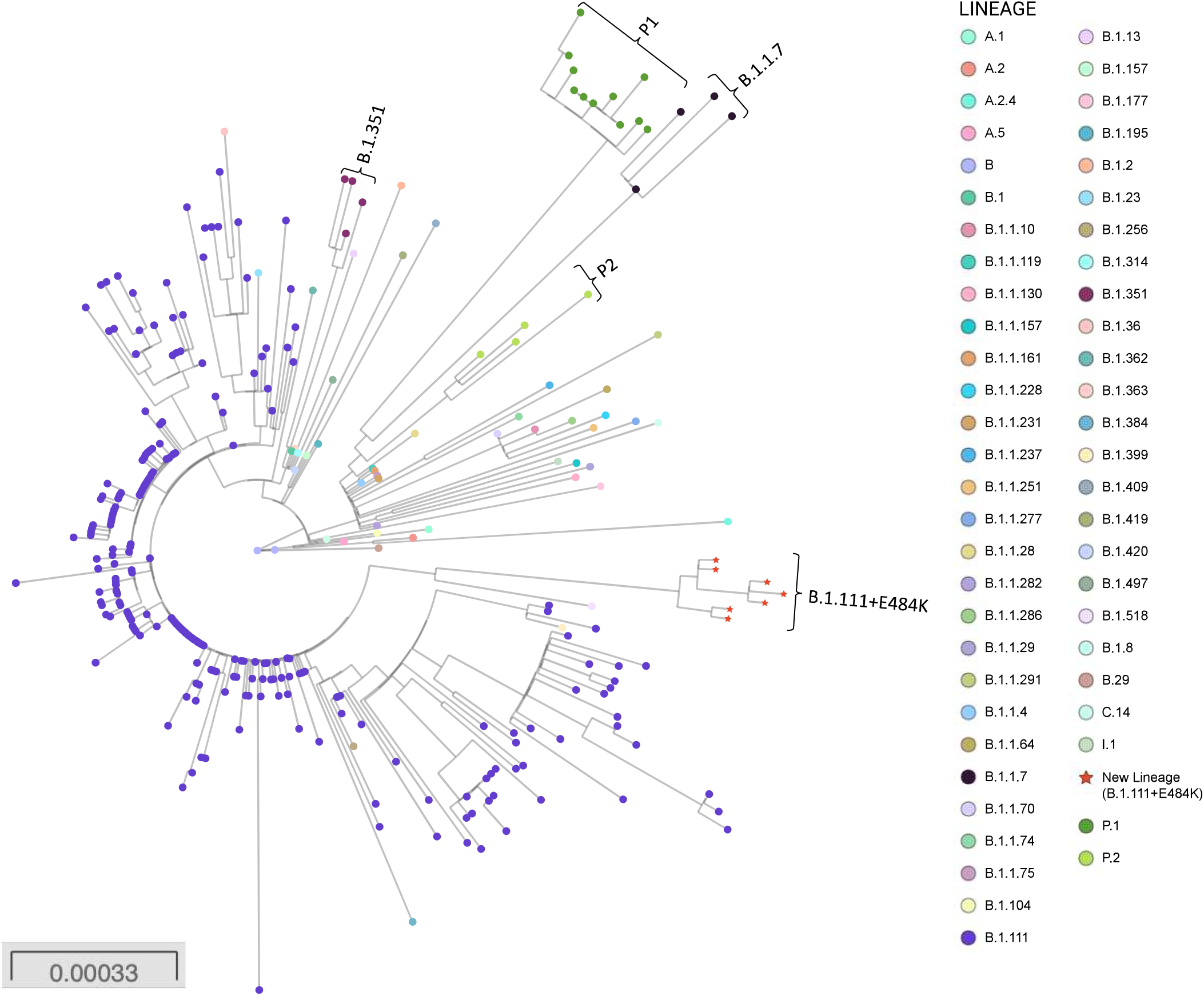
Phylogenetic tree of the new lineage of SARS-CoV-2 emerging from B.1.111 lineage. The tree was reconstructed by maximum likelihood with the estimated GTR+F+I nucleotide substitution model for the dataset of 304 full-length genomes, representative of the principal recently emerging lineages. The interactive tree can be accessed in the following link: https://microreact.org/project/nFBT2K1JdjcMuEPH7u32Ar/6d9eb0c0. Red stars represent the sequences belonging to the new lineage.

The large list of distinctive mutations at the nucleotide and protein levels (Table 1) are consistent with the existence of a common recent ancestor for the Colombian sequences and three other reported sequences from USA (2 sequences) and Belgium (1 sequence). The intra-lineage and between-lineages p-distances suggest a drastic divergence of the new lineage from the most closely related lineages (Appendix 2 Table S1). While increasing the sample size could help to reconstruct the gradual accumulation of mutations leading to divergence from the B.1.111 ancestor, a plausible explanation for the origin of this new lineage could also be the existence of a strong selection pressure on the virus population in an unknown context (e.g., natural infection in a population reaching herd immunity, convalescent plasma or monoclonal antibodies treatment, chronic infection in immunocompromised patients, replication in a different vertebrate species, etc.) (5,7–9). The result of the analysis by IFEL and MEME (using a *P*-value threshold of □ ≤0.3) is suggestive of positive selection in 7 codons related to some distinctive non-synonymous substitutions (Appendix 2 Table S2).

**Table 1.** The E484K substitution in the Spike protein has emerged independently in several lineages and countries where they were first reported.

|  |  |  | Country of |  |  |
| --- | --- | --- | --- | --- | --- |
| Lineage | Country of<br>first reported | Earliest<br>collection | Lineage | first<br>reported | Earliest<br>collection |
| + 484K | seq | date | + 484K | seq | date |
| A | Nigeria | 2021-01-07 | B.1.139 | USA | 2020-05-22 |
| A.2 | Spain | 2020-03-31 | B.1.146 | Brazil | 2021-01-11 |
| A.23.1 | England | 2020-12-26 | B.1.160 | Netherlands | 2020-11-07 |
| B.1 | Switzerland | 2020-03-21 | B.1.165 | England | 2021-01-08 |
| B.1.1 | USA | 2020-12-09 | B.1.177 | Switzerland | 2020-09-30 |
| B.1.1.1 | England | 2020-06-06 | B.1.177.4 | England | 2020-11-14 |
| B.1.1.10 | England | 2020-12-26 | B.1.2 | Canada | 2020-12-24 |
| B.1.1.103 | Bangladesh | 2020-12-19 | B.1.214 | Belgium | 2021-01-28 |
| B.1.1.105 | USA | 2021-01-04 | B.1.234 | USA | 2021-01-14 |
| B.1.1.130 | Russia | 2020-11-17 | B.1.235 | Nigeria | 2021-01-08 |
| B.1.1.145 | South Africa | 2020-10-09 | B.1.237 | South Africa | 2020-09-16 |
| B.1.1.163 | Israel | 2020-12-24 | B.1.241 | USA | 2020-07-08 |
| B.1.1.164 | Australia | 2020-04-16 | B.1.243 | USA | 2021-01-18 |
| B.1.1.184 | Russia | 2020-11-09 | B.1.260 | USA | 2021-01-06 |
| B.1.1.207 | USA | 2020-11-18 | B.1.274 | Kuwait | 2020-12-27 |
| B.1.1.220 | USA | 2020-12-31 | B.1.279 | Canada | 2020-04-20 |
| B.1.1.235 | England | 2020-08-17 | B.1.280 | USA | 2021-01-14 |
| B.1.1.269 | Belgium | 2020-11-10 | B.1.281 | Bahrain | 2020-11-09 |
| B.1.1.273 | South Africa | 2020-10-05 | B.1.311 | USA | 2021-01-15 |
| B.1.1.28 | England | 2020-12-08 | B.1.316 | England | 2020-11-14 |
| B.1.1.29 | USA | 2020-12-28 | B.1.351 | South Africa | 2020-10-08 |
| B.1.1.305 | USA | 2021-02-04 | B.1.369 | USA | 2020-10-06 |
| B.1.1.306 | India | 2020-09-01 | B.1.400 | USA | 2020-12-10 |
| B.1.1.316 | Bangladesh | 2021-01-19 | B.1.416 | Gambia | 2020-09-22 |
| B.1.1.33 | Brazil | 2020-11-11 | B.1.441 | Egypt | 2021-01-03 |
| B.1.1.34 | South Africa | 2020-10-06 | B.1.486 | USA | 2020-10-22 |
| B.1.1.57 | South Africa | 2020-10-09 | B.1.487 | USA | 2021-01-15 |
| B.1.1.67 | Singapore | 2021-01-22 | B.1.525 | England | 2020-12-15 |
| B.1.1.7 | England | 2020-12-17 | B.1.526 | USA | 2020-12-16 |
| B.1.1.74 | England | 2020-04-30 | C.1 | South Africa | 2020-11-10 |
| B.1.1.85 | USA | 2021-01-08 | D.2 | Australia | 2020-10-27 |
| B.1.110.3 | USA | 2020-09-01 | H.1 | South Africa | 2020-12-21 |
| **B.1.111** | **Colombia** | **2020-12-26** | P.1 | Brazil | 2020-12-04 |
| B.1.134 | USA | 2020-08-25 | P.2 | Brazil | 2020-04-15 |

<sup>1</sup> Analysis performed for the sequences available up to February 14, 2021 in GISAID.

The presence of E484K located at the receptor binding domain (RBD) is of special relevance as it has been associated to the phenotypic properties of two well described lineages, B.1.1.28 and B.1.351 and it is being suggested to be responsible for a considerably lower neutralizing activity in vitro from convalescent plasma (8,10,11), although the cell-mediated immunity could not be affected by the distinctive mutations (12). In the same way, S249L is located at the N-terminal domain (NTD), the second domain most frequently targeted by neutralizing antibodies (11). The potential impact of other changes in critical proteins for viral replication (e.g., Helicase, 2’-O-ribose methyltransferase, etc.) is to be determined.

Despite increasing effort in the routine genomic surveillance in Colombia, the new lineage has only been detected from samples collected during late December to mid-January mainly from the Caribbean region of the country, which supposes a major effort is necessary to determine the epidemiologic contribution in the different cities.

An obligatory question that arises from the current analysis of the novel lineage and the evidence of 68 lineages with the evolutionary convergence at the Spike amino acid E484K is related to the context of the emergence of highly divergent lineages, and the selection of specific substitutions. The fact that some amino acid changes have appeared independently in these lineages is not plausibly explained by chance, but probably by the result of a selective immune pressure. Many hypotheses have been raised without conclusive support. One of them is related to the chronic infection in immunocompromised patients and the administration of under-neutralizing antibody titers during convalescent plasma or monoclonal antibody therapies (7,13–15).

In the context of pandemic spread of the virus, an enormous virus population size is expected, as it is also the emergence of virus variants that could also make possible the emergence of antibody-resistant mutants in the context of natural infection in immunocompetent people. It is necessary to evaluate the impact of this genetic background in the neutralization efficacy of convalescent sera/plasma from acquired immunity.

## Ethics statement

In the submission system, your current ethics declaration states: “The INS is the reference lab and health authority of the national network of laboratories. In case of a public health emergency or when scientific research in public health is required, the national law 9/1979, decrees 786/1990 and 2323/2006, authorizes the INS to use the biological material and associated epidemiological information without informed consent, including the anonymous disclosure of results. This study was approved in the project CEMIN-4-2020 by the Ethical commit of the Instituto Nacional de Salud Colombia and performed following the ethical standards noted in the 1964 Declaration of Helsinki and its later amendments. The information used for this study comes from secondary sources of data that were anonymized and do not represent a risk to the community. “

## Data Availability

The authors approved availability of all data referred to in the manuscript and web links.

## Acknowledgments

The authors thank the National Laboratory Network for routine virologic surveillance of SARS-CoV-2 in Colombia. We also thank all researchers who deposited genomes in GISAID’s EpiCoV Database contributing to genomic diversity and phylogenetic relationship of SARS-CoV-2. We thank Rotary International and Charlie Rut Castro for equipment’s donation. Finally, we thank red RENATA and Universidad Industrial de Santander for the bioinformatic assistance.

This work was funded by the Project CEMIN-4-2020 Instituto Nacional de Salud. The funders had no role in study design, data collection and analysis, decision to publish, or preparation of the manuscript.

## Address for correspondence

